# Alterations of the gut microbiome are associated with epigenetic age acceleration and physical fitness

**DOI:** 10.1101/2023.07.05.23292191

**Authors:** Ferenc Torma, Csaba Kerepesi, Matyas Jokai, Peter Bakonyi, Erika Koltai, Gergely Babszki, Balazs Ligeti, Regina Kalcsevszki, Kristen M. McGreevy, Steve Horvath, Zsolt Radak

**Author notes:** Correspondig author: Zsolt RADAK, PhD., DSc. Joint first-authors (Ferenc Torma, Csaba Kerepesi).

## Abstract

Epigenetic clocks can measure aging and predict the incidence of diseases and mortality. Higher levels of physical fitness are associated with a slower aging process and a healthier lifespan. Microbiome alterations occur in various diseases and during the aging process, yet their relation to epigenetic clocks is not explored. To fill this gap, we collected metagenomic, epigenetic and exercise-related data from physically fit individuals and applying epigenetic clocks, we examined the relationship between gut flora, epigenetic age acceleration and physical fitness. We revealed that an increased entropy in the gut microbiome is associated with accelerated epigenetic aging, lower fitness or impaired health status. We also observed that, in general, accelerated epigenetic aging can be linked to the abundance of pro-inflammatory and other pathogenic bacteria and decelerated epigenetic aging or high fitness level can be linked to the abundance of anti-inflammatory bacteria. Overall our data suggest that alterations in the microbiome can be associated with epigenetic age acceleration and physical fitness.

## Introduction

Currently, there are very limited options for increasing the maximum lifespan in humans. On the other hand, a substantial body of evidence, from several studies^1, 2^, has consistently shown that a healthy lifestyle can increase the expected lifespan and reduce the incidence of lifestyle-related diseases. DNA methylation-based aging clocks (shortly epigenetic clocks) are developed to measure biological age that may be largely influenced by lifestyle among other environmental and genetic factors^3, 4^. The first-generation epigenetic clocks (such as Horvath’s pan-tissue clock, the blood-based Hannum clock and the Skin and Blood clock) can predict age accurately and exhibit associations with clinical biomarkers and mortality risk^5–7^. Second-generation epigenetic clocks, such as PhenoAge, GrimAge and DunedinPACE show even stronger associations with mortality risk and some age-related conditions^8–10^. Very recently we developed DNAmFitAge which is based on genes that are related to physical fitness^11^. It was shown that physically fit people have a younger DNAmFitAge and experience better age-related outcomes: lower mortality risk, coronary heart disease risk, and increased disease-free status. DNA methylation-based aging clocks are plastic and readily respond to lifestyle modifications and stress^12, 13^. Another very plastic system in the human body is the microbiome, which is characterized by rapid change in childhood, followed by relatively long lifestyle-associated stability, and finally age-associated modification^14^. The results of a recent study using the microbiome profile of 9000 subjects revealed that with aging the microbiome flora is getting more and more unique, which is associated with immune regulation, inflammation, and longevity, moreover, over 85 years the high relative abundance of Bacteroides and having a low gut microbiome uniqueness measure were both associated with significantly decreased survival in the course of 4-year follow-up^14^. The microbiota of the gut is crucial for breaking down dietary nutrients, regulating intestinal and systemic immune responses, producing small molecules critical for intestinal metabolism, and generating several gasses that can modulate cellular function. Due to the complex function of the gut microbiome, the diversity of microbes can be defined as the range of different kinds of unicellular organisms, bacteria, archaea, protists, and fungi^15^. However, the possible role or connection of the microbiome to epigenetic aging and physical fitness is still unknown. Recently attempts were made to create a pipeline to study the association of the architecture of microbiome and host diseases^16^ or developing aging clocks based on taxonomic and functional profiling^17, 18^. Moreover, it appears that the well-known difference in the mean lifespan of females and males could be associated with measurable differences in the microbiome and this sexual dimorphism in the microbiome (i.e. microgenderome) has high relevance to disease susceptibility^19^. These approaches suggest the great potential of metagenomic investigations on human health, diseases, and aging. Here we examine associations between microbiome flora, DNA methylation-based aging, and the level of physical fitness.

## Results

### Characterisation of the gut microbiome of physically fit males and females

We collected metagenomic, epigenetic and exercise-related data from 80 master athletes with ages between 38 and 84 years and applying epigenetic clocks, we examined the relationship among the microbiome, epigenetic age acceleration and physical fitness (Fig. 1A). Females and older individuals are slightly overrepresented in our volunteer cohort (Fig. 1B). First, we characterised the gut microbiome of physically fit males and females in the phylum level. *Firmicutes* was the highest abundant taxa overall (mean: 0.556, std: 0.223) followed by *Bacteriodetes* (mean: 0.172, std: 0.223). We observed a remarkable difference in phylum and species distribution by gender (Fig. 1C, Extended Data Fig 1., respectively). *Firmicutes* had a significantly lower relative abundance in females compared to males, while the relative abundance of *Proteobacteria* was significantly higher in females (Fig. 1D). As males and females of our cohort express remarkable differences regarding the number of samples, microbiome composition, epigenetic age acceleration and exercise-related parameters, we conduct data analysis separated by gender.

**Fig. 1.**
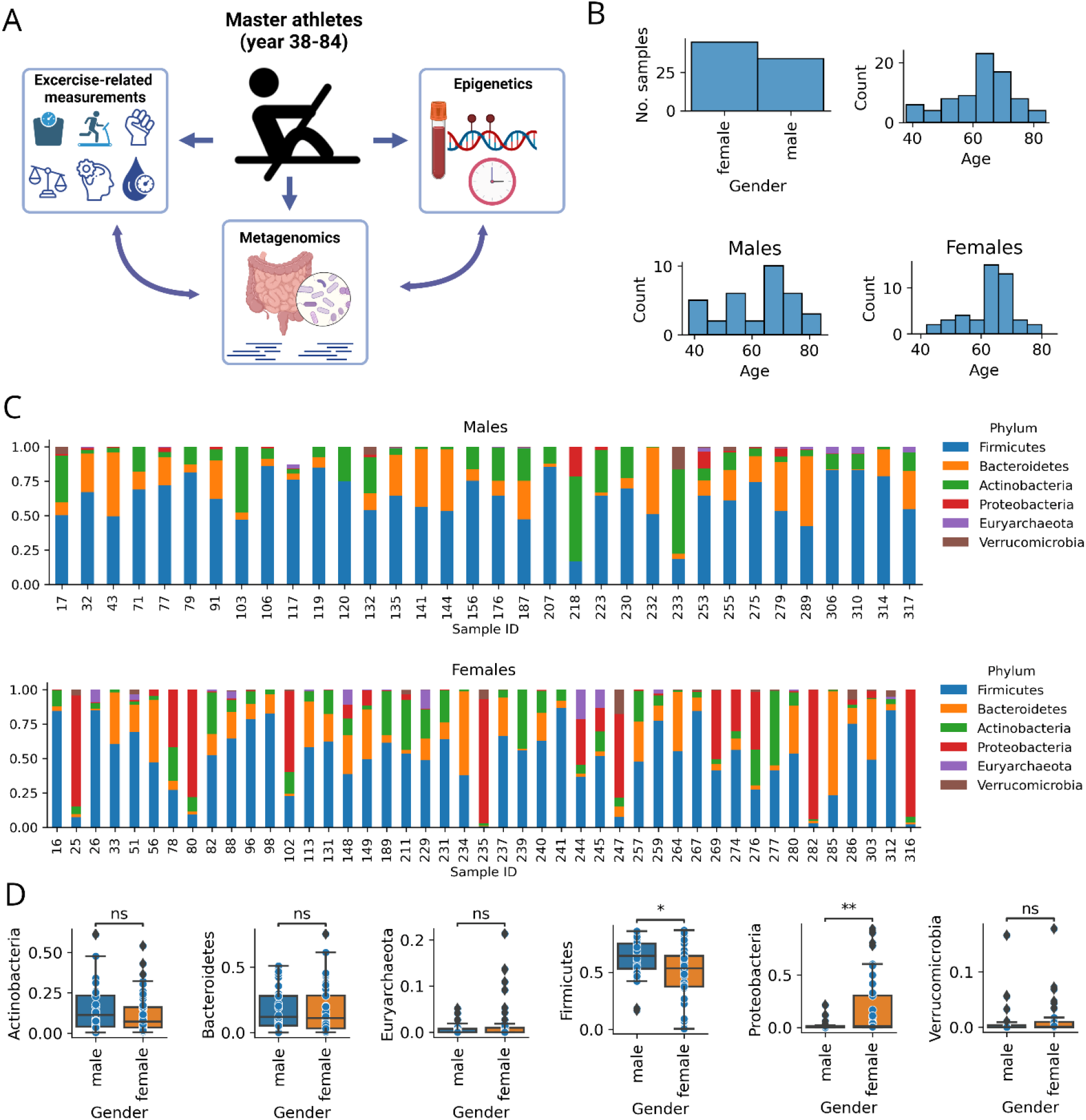
Characterisation of the gut microbiome of physically fit males and females. **(A)** Overview of the study. We collected metagenomics, epigenetics and exercise-related data from 80 physically fit individuals with age between 38 and 84 years, and applying epigenetic clocks, we examined the relationship of epigenetic age acceleration, gut flora and physical fitness. **(B)** Gender and age distribution of the study samples. **(C)** Phylum distributions for males and females, separately (only the abundant phyla are displayed). **(D)** Differences of mean relative abundances between males and females separated by phyla.

### Associations of the microbiome and epigenetic aging clocks

We investigated the relationships between epigenetic aging clocks and the results of shotgun sequencing of the microbiome (Fig. 2, Extended Data Fig. S2). We calculated the diversity of the microbiome (measured by Shannon entropy of the relative abundances) at the seven taxonomic levels. We found significant positive correlations between age acceleration (i.e. the advanced biological age of an individual) and microbiome diversity for some epigenetic clocks at some taxonomic levels (four cases for males and one case for females) (Fig. 2AB). In general, our data suggests that an increased entropy in the gut microbiome may be associated with accelerated epigenetic aging.

**Fig. 2.**
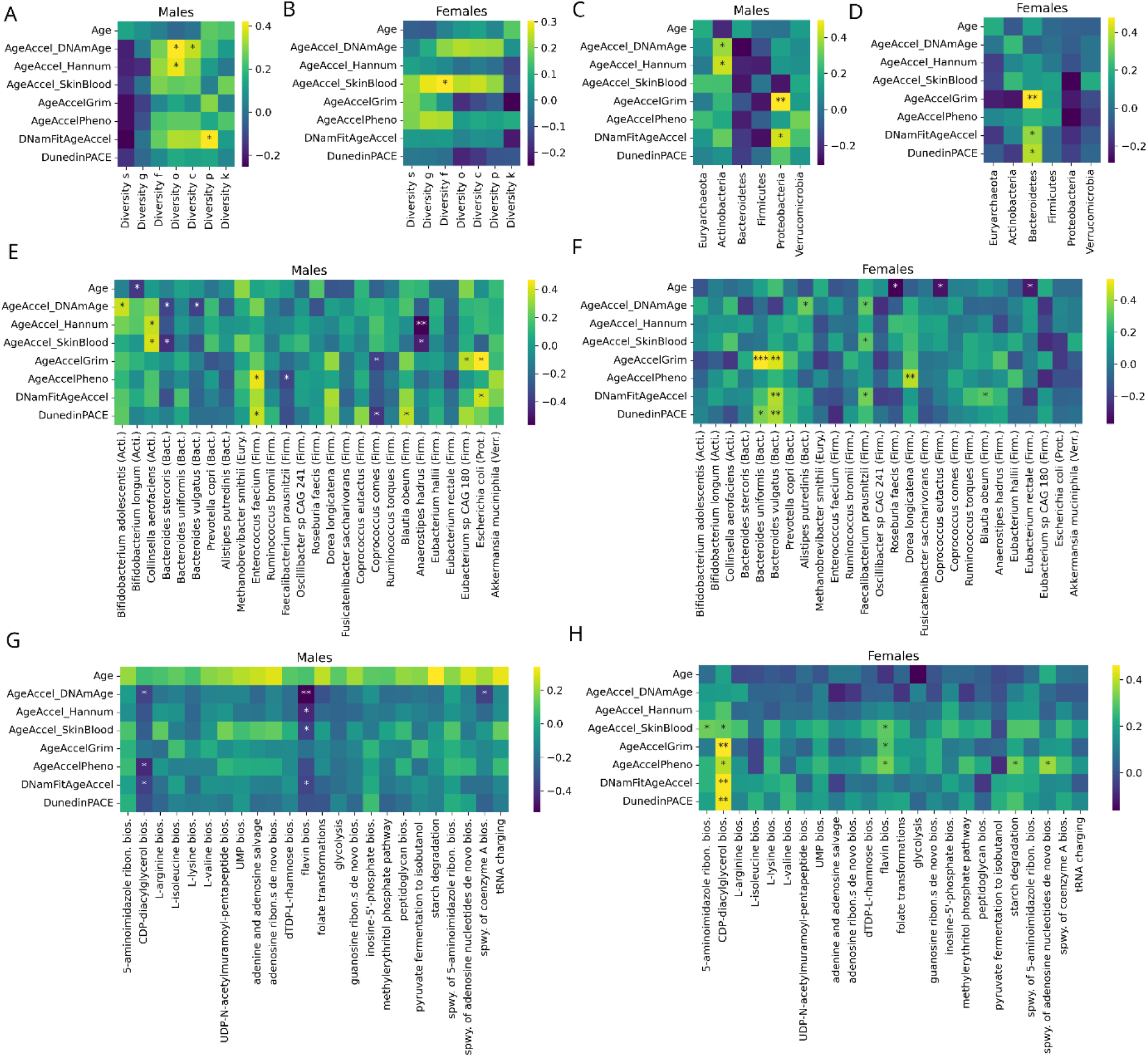
Associations of the gut microbiome and epigenetic aging clocks. **(AB)** Correlations between the diversity of the microbiome (measured by Shannon entropy of the relative abundances) at the level of species (s), genus (g), family (f), order (o), class (c), phylum (p) and kingdom (k) and chronological age, age accelerations as well as pace of aging. **(CD)** Similar correlation analysis for the relative abundance of phyla. **(EF)** Similar correlation analysis for the abundant species of the gut microbiome. Phyla are also displayed in parentheses abbreviated by the first four letters of the phylum. **(GH)** Similar correlation analysis for the most abundant bacterial pathways in the gut microbiome (spwy., superpathway; bios., biosynthesis).

For males, at the phylum level, the relative abundance of *Actinobacteria* and *Proteobacteria* both positively correlated with the age acceleration calculated by two-two epigenetic clocks (Horvath’s pan-tissue DNAm age and the Hannum clock as well as DNAmFitAge and GrimAge, respectively) (Fig. 2C). For females, at the phylum level, the relative abundance of *Bacteroidetes* positively correlated with the age acceleration by three epigenetic clocks (DNAmFitAge, GrimAge, and DunedinPACE) (Fig. 2D).

To refine the above results at the species level, we investigated the relationship between epigenetic clocks and the relative abundance of the abundant species. We considered only strong associations (agreed by at least two epigenetic clocks and/or p-value smaller than 0.01). In males, the positive association of *Actinobacteria* and age acceleration seems to be driven by *Bifidobacterium adolescentis*, *Collinsella aerofaciens*, while the positive association of *Proteobacteria* can be linked to *Escherichia coli* (Fig. 2E). Other remarkable positive associations with age acceleration or pace of aging were observed in the case of *Enterococcus faecium*. Interestingly, strong negative associations with age acceleration also were found in the case of three species: *Bacteroides stercoris, Coprococcus comes* and *Anaerostipes hadrus*. In females, the positive association of *Bacteroidetes* phylum and age acceleration is clearly linked to two species, *Bacteroides uniformis*, and *Bacteroides vulgatus* (Fig. 2F). However, other remarkable positive associations with age acceleration were observed in two *Firmicutes* species: *Faecalibacterium prausnitzii* and *Dorea longicatena*.

We also examined the relationship between bacterial pathways of the microbiome and epigenetic clocks. CDP-diacylglycerol biosynthesis and flavin biosynthesis showed negative correlations with age acceleration and pace of aging in males (Fig. 2G). Interestingly, the same pathways showed positive correlations with age acceleration and pace of aging in females (Fig. 2H).

### Associations of the microbiome and exercise-related parameters

We also investigated the relationship between exercise-related measurements, relative abundance, and bacterial species- catalyzed pathways (Fig. 3, Extended Data Fig. 4). In males, we found significant negative correlations between microbial diversity and the parameters VO2max, JumpMax, and Redox Balance and positive correlations with trygliceride levels (Fig. 3A). In females we found significant negative correlations between microbial diversity and BMI and cognitive test performance (Fig. 3B). Overall, our data suggest that an increased entropy in the gut microbiome may be associated with low fitness or impaired health status.

**Fig. 3.**
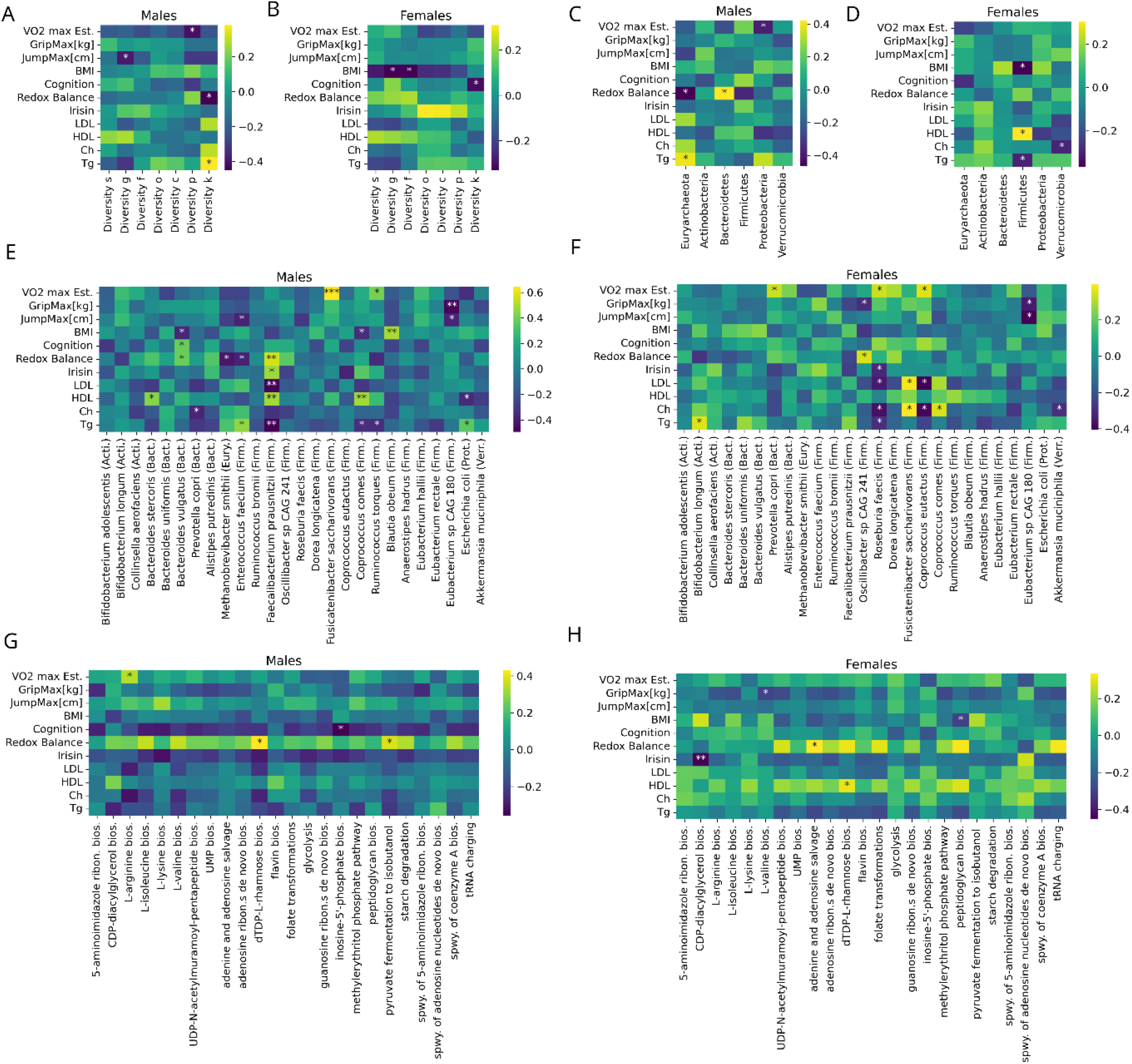
Associations of the microbiome and exercise-related measurements. **(AB)** Correlations between the diversity of the microbiome (measured by Shannon entropy of the relative abundances) at the level of species (s), genus (g), family (f), order (o), class (c), phylum (p) and kingdom (k) and exercise-related parameters. **(CD)** Similar correlation analysis for the relative abundance of phyla. **(EF)** Similar correlation analysis for the abundant species of the gut microbiome. Phyla are also displayed in parentheses abbreviated by the first four letters of the phylum. **(GH)** Similar correlation analysis for the most abundant bacterial pathways in the gut microbiome (spwy., superpathway; bios., biosynthesis).

In males, at the phylum level, our statistical analysis revealed a positive relationship between *Euryarchaeota* and triglyceride levels, as well as, *Bacteroidetes* and redox balance, while negative associations were observed between *Euryarchaeota* and redox balance, as well as, *Proteobacteria* and VO2max (Fig. 3C). In females, *Firmicutes* were negatively associated with BMI and triglyceride levels and positively associated with HDL levels (Fig. 3D). Furtheremore, the greater relative abundance of *Verrucomicrobiota* was associated with lower levels of cholesterol.

We refined the above analysis at the species level and focused on the highly significant associations (p-value smaller than 0.01) or the cases when consistency of multiple associations appeared. In males, VO2max showed a remarkable high positive correlation with the abundance of *Fusicatenibacter saccharivorans* (Fig. 3E). *Faecalibacterium prausnitzii* showed a high positive correlation with redox balance and HDL, and at the same time, low correlations with LDL and triglyceride level. *Coprococcus comes* relative abundance expressed a positive association with HDL levels, while *Blautia obeum* showed a positive association with BMI. The genus *Eubacterium*, which consists of Gram-positive bacteria, exhibited a negative relationship with GripMax and JumpMax.

In females, the abundance of *Roseburia faecis* showed a negative relationship to irisin, LDL, triglyceride, and cholesterol levels (Fig. 3F). The genus *Eubacterium* exhibited a negative relationship with GripMax and JumpMax, similarly to males.

We examined the relationship between physical fitness-related biomarkers and the molecular pathways that are catalyzed by bacterial species in the microbiome (Fig. 2E). Redox balance showed a positive association with *dTDP-L-rhamnose biosynthesis* and *pyruvate fermentation to isobutanol* in males as well as with *adenine and adenosine salvage* pathway in females. Another remarkable observation is that, in females, irisin levels showed a low correlation to cdp-diacylglycerol pathway.

## Discussion

The microbiome is highly plastic to acute nutritional and exercise interventions, or health status and relatively stable because longitudinal studies suggest that the composition of intestinal microbiota does not drastically change in adults within the periods examined^20^.

Males and females have differences in lifespan, sex hormones, muscle mass, VO2max and the power of the immune system^19, 21^, therefore our observation on the difference of male and female microbiome could be expected. Early results of the Human Microbiome Project revealed that the highest abundant phyla in the gut microbiome of the general healthy population are Firmicutes and Bacteriodetes^22^. Consistently, these two cohorts were the most abundant in our highly fit master athlete cohort, however, the proportion of the two phyla is the opposite in our master athlete cohort compared to the general healthy population.

The *Proteobacteria* phylum contains a number of pathogens such as *Brucella*, *Rickettsia*, *Shigella*, *Salmonella*, *Yersinia*, *Helicobacter* and *Escherichia*. Therefore it was suggested that the increased prevalence of *Proteobacteria* is a potential diagnostic signature of dysbiosis and risk of disease^23^. We found a positive correlation between *Proteobacteria* and age acceleration in males with a species level dominance of *Eschericia coli*. Therefore it is possible that the infections of some pathogen *Eschericia coli* strain accelerate epigenetic aging. This would be consistent with previous findings where accelerated epigenetic aging was reported in the case of other pathogen infection such as HIV and SARS-CoV-2^13, 24, 25^.

A correlation between *Bifidobacterium adolescentis* and mental disorders such as depression and anxiety was reported^26^. *Collinsella aerofaciens* is identified as pro-inflammatory bacteria in rheumatoid arthritis, psoriasis, Crohn’s disease, inflammatory bowel disease, atherosclerosis, non-alcoholic steatohepatitis, type 2 diabetes, and COVID-19^27^. Both of these species are associated with accelerated epigenetic aging in males according to our findings.

We observed a negative correlation between epigenetic age acceleration and the relative abundance of *Anaerostipes hadrus,* which has been identified as an anti-inflammatory, butyrate-producing species, and butyrate has been suggested to beneficially affect gut health^28^. In summary, our findings consistently show accelerated aging linked to pro-inflammatory and pathogenic bacteria and decelerated aging linked to anti-inflammatory bacteria.

Bacteria relative abundaces that strongly correlated with age accelerations did not show significant correlations with choronological age. This suggest that age acceleration (i.e advanced age) does not cause an increase in the abundance of pro-inflammatory bacteria but rather an increase in the abundance of pro-inflammatory bacteria cause age acceleration. With the same argument, anti-intaflammatory bacteria may cause age deceleration.

In females, we also observed a remarkable positive correlation with age acceleration for the relative abundance of *Dorea longicatena*, which is linked to metabolic risk markers in obesity^29^, and its level is higher in over-weight subjects^30^. Epigenetic age acceleration was linked to obesity in previous studies^3, 8^. In females, we also observed a positive association between *Bacteroidetes* pyhlum and age acceleration that was clearly linked to two species: *Bacteroides uniformis*, and *Bacteroides vulgatus*. A gut microbiome analysis linked *Bacteroides vulgatus* with ulcerative colitis severity^31^.

*Fusicatenibacter saccharivorans* highly correlated with VO2max in males, and this gram-stain-positive bacteria reportedly reduced inflammation^32^ and contribute to the production of short-chain fatty acids (SCFA)^33^, which are crucial to the regulation of tight junction proteins involved in the permeability of the epithelial barrier in the colon that are associated with obesity and insulin resistance^34^. Through receptor activation, gut-derived SCFA is an active player in signaling and metabolic organ-to-organ communication. However, it is not known why *Fusicatenibacter saccharivorans* level is positively linked to cardiovascular fitness. One possible mechanism could be that *Fusicatenibacter* produces butyrate which supplementation can activate peroxisome proliferator-activated receptor-gamma coactivator-1alpha levels and the activities of AMP kinase and p38 in skeletal muscle of mice^35^. Higher mitochondrial content can readily lead to increased VO2max, however, it is not known whether a similar mechanism could happen in humans.

The genus *Eubacterium sp. CAG 180* exhibited a negative relationship with GripMax and JumpMax in both genders, but there are no associations for other abundant *Eubacteria* species (*Eubacterium rectale* and *Eubacterium hallii*). While the greater abundance of some *Eubacteria* species suggested having health benefits (*Eubacterium ventriosum*, *Eubacterium ventriosum*)^37^ there is no report for *Eubacterium sp. CAG 180*.

Redox balance showed a positive association with *dTDP-L-rhamnose biosynthesis* and *pyruvate fermentation to isobutanol* in males as well as with *adenine and adenosine salvage* pathway in females. This observation suggests that redox balance in a cell has a positive effect on the above-mentioned molecular pathways, potentially indicating a regulatory role of redox balance in various cellular and metabolic pathways. VO2max was correlated with L-arginine biosynthesis in males, which fits well with the result of the study which reported increased VO2max after L-arginine supplementation^38^.

It was also observed that irisin levels negatively related to cdp-diacylglycerol pathway in females. The cdp-diacylglycerol pathway is involved in the synthesis of phosphatidylcholine, a major component of cell membranes. A negative correlation between irisin levels and this pathway suggests that higher levels of irisin may be associated with lower activity or expression of genes involved in the cdp-diacylglycerol pathway. These correlations indicate potential connections between cognitive performance, hormone levels, and specific metabolic pathways.

## Methods

### Study Population and Physiology Tests

This study was approved by the Institutional Ethical Review Board and National Science and Research Ethical Committee (https://ett.aeek.hu/tukeb/), Hungary in accordance with the Helsinki Declaration and the regulations applicable in Hungary (25167-6/2019/EUIG).

Eighty volunteers were tested, and based on the results of Chester’s step, the maximal oxygen uptake was calculated and used as a measure of cardiovascular fitness. Maximum handgrip force is often used to measure the age-associated decline in overall muscle strength. The dynamic strength of the legs was assessed by measuring the maximum vertical jump using a linear encoder. Body mass index was determined using the body composition monitor BF214 (Omron, Japan).

### Measurement of Irisin

Plasma irisin levels were quantified using commercially available ELISA kits (EK-067–29, Irisin Recombinant, Phoenix Pharmaceuticals, Inc, Burlingame, USA). All samples from each subject were analyzed using the same plate (intra-assay). The coefficients of variation for intra-assay and inter-assay were 4.1% and 15.2%, respectively.

### Assessment of Redox Balance

Redox balance was calculated as the ratio of biological antioxidant power (BAP) to derivatives of reactive oxygen metabolites (d-ROMs). BAP was measured by mixing ferric chloride with thiocyanate derivative in blood plasma samples. After incubation, the reduction of ferric ions was measured at 505 nm. BAP assays were performed using a FREE Carpe Diem analyzer. The total amount of organic hydroperoxides in blood was estimated using the d-ROMs test, as described previously^39^.

### Microbiome Assay

Stool samples were collected for analysis of gut microbiota. Participants were provided with instructions on proper methods for stool collection, and all necessary materials were included in a convenient specimen collection kit. The samples were stored at - 80 °C until further analysis. A frozen aliquot (200 mg) of each fecal sample was suspended in 250 ml of guanidine thiocyanate, 0.1 M Tris, pH 7.5, and 40 ml of 10% N-lauroyl sarcosine. DNA extraction was then performed as previously described^40^, and the DNA concentration and molecular size were estimated using a nanodrop (Thermo Scientific) and agarose gel electrophoresis.

### Illumina Sequencing

Fecal DNA was used as input for the Illumina Nextera® XT DNA Sample Preparation Kit to construct indexed paired-end DNA libraries, following the previously described method^41^. DNA library preparation followed the manufacturer’s instructions (Illumina). The workflow indicated by the provider was used for cluster generation, template hybridization, isothermal amplification, linearization, blocking and denaturing, and hybridization of the sequencing primers. The base-calling pipeline (version IlluminaPipeline- 0.3) was used to process the raw fluorescent images and call sequences. One library (clone insert size 200 base pairs (bp)) was constructed for each of the first batch of 15 samples, two libraries with different clone insert sizes (135 and 400 bp) were constructed for each of the second batch of 70 samples, and one library (350 bp) was constructed for each of the third batch of 207 samples.

### Bioinformatics Analysis

The quality of raw and trimmed reads was assessed using FastQC and MultiQC. Low-quality sequences were filtered and trimmed using Trimmomatic, discarding sequences with a minimum length of 36 and low-quality base calls (phred score < 30). Reads aligning to the human reference genome (GRCh38) were removed to eliminate host contamination (using bowtie2 v2.4.2). Taxonomic characterization was performed using MetaPhlAn3, and pathway abundance and other molecular function profiles (such as GO) were estimated using the HUMAnN3 pipeline. We discarded a taxon if its average abundance over the 80 samples is less than 1%.

### Measurement of DNA Methylation

Epigenome-wide DNA methylation was measured using the Infinium MethylationEPIC BeadChip (Illumina Inc., San Diego, CA) following the manufacturer’s protocol. Briefly, 500 ng of genomic DNA was bisulfite-converted using the EZ-96 DNA Methylation MagPrep Kit (Zymo Research, Irvine, CA, USA) with the KingFisher Flex robot (Thermo Fisher Scientific, Breda, Netherlands). The samples were plated in a randomized order. Bisulfite conversion was performed according to the manufacturer’s protocol with the following modifications: 15 µl MagBinding Beads were used for DNA binding, and the conversion reagent incubation was carried out in a cycle protocol of 16 cycles at 95°C for 30 seconds followed by 50°C for 1 hour. After the cycle protocol, the DNA was incubated for ten minutes at 4°C. Next, DNA samples were hybridized on the Infinium MethylationEPIC BeadChip (Illumina Inc., San Diego, CA) using 8 µl of bisulfite-treated DNA as the starting material.

### Quality Control of the DNA Methylation Data

Quality control of the DNA methylation data was performed using the minfi, Meffil, and ewastools packages in R version 4.0.0. Samples that failed technical controls, including extension, hybridization, and bisulfite conversion according to Illumina’s criteria, were excluded. Samples with a call rate < 96% or with at least 4% of undetected probes were also excluded. Probes with a detection p-value > 0.01 in at least 10% of the samples were considered undetected and excluded. Probes with a bead number < 3 in at least 10% of the samples were also excluded. The “noob” normalization method in R was used to quantify methylation levels.

### DNA methylation (or epigenetic) aging clocks

Aging clocks were applied using Horvath’s online age calculator (https://dnamage.genetics.ucla.edu/) and the DunedinPACE R package (https://github.com/danbelsky/DunedinPACE). We applied the Horvath pan-tissue clock^5^, the blood-based Hannum clock^6^, Skin and Blood clock^7^, PhenoAge clock^8^, GrimAge clock^9^, DNAmFitAge clock^11^ and the DunedinPACE clock^10^. We calculated age acceleration as the residual, per sample, after fitting predicted ages to chronological ages^42^ (i.e. age acceleration is the deviation from the trend) (Extended Data Fig. 2A-F). As expected, epigenetic clocks highly correlated with age and with each other in our cohort (Extended Data Fig. 2A-H) and age accelerations were independent of age (Extended Data Fig. 2I).

### Statistical analysis

We used the Python packages for statistical analysis. Two-sided t-test was calculated for comparing two groups. If p values were indicated by an asterisk, we used the notations as follows: ns, p > 0.05; *, 0.01 < p ≤ 0.05; **, 0.001 < p ≤ 0.01; ***, p ≤ 0.001. Correlations were evaluated by Pearson correlation coefficient (r).

## Data Availability

All data produced in the present study are available upon reasonable request to the authors

https://www.tf.hu

## Founding

CK was supported by the European Union project RRF-2.3.1-21-2022-00004 within the framework of the Artificial Intelligence National Laboratory, Hungary. ZR acknowledge support from the National Excellence Program (126823) National Science and Research Found (OTKA142192) and Scientific Excellence Program TKP2021-EGA-37 at the Hungarian University Sport Science, Innovation and Technology Ministry, Hungary, as well as Post-Covid 2021-28 grant by National Academy of Science, Hungary.

## Authors’ contributions

Conceptualization, Z.R. Methodology, F.T. C.K. S.H. K.M and Z.R; Investigation, M.J., F.T., E.K., G.B., P.B., and Z.R. Formal Analysis, K.M., F.T., R.K. C.K., and S.H. Writing – Original Draft, Z.R., and C.K. Writing – Review & Editing, C.K. S.H. B.S. and Z.R. Supervision, Z.R.

## SUPPLEMENTARY FIGURES

**Extended Data Fig. 1.**
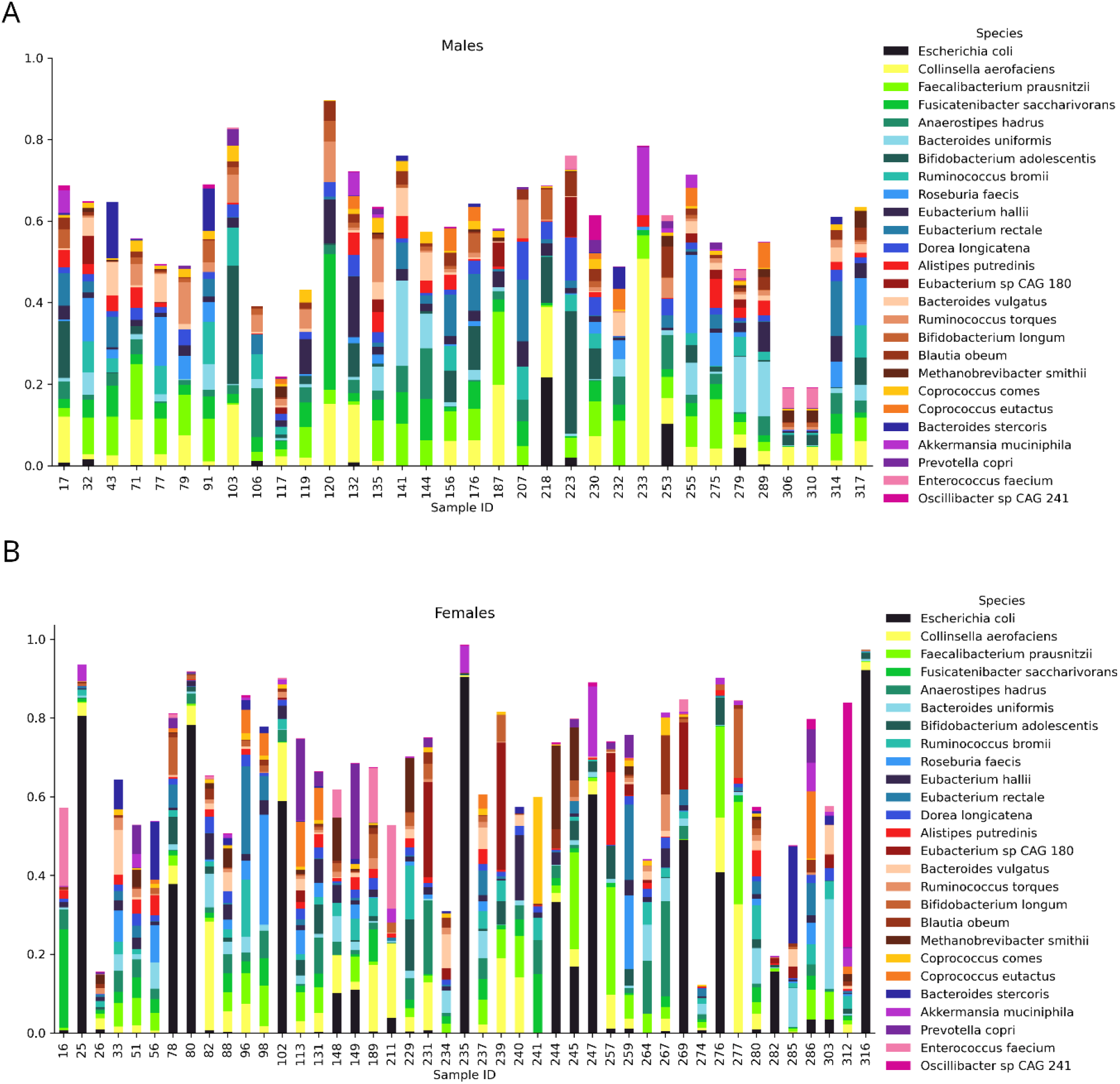
Species distributions of the gut microbiome of our cohort. **(A)** males **(B)** females. Only the abudant species are displayed. This figure is related to Fig. 1C.

**Extended Data Fig. 2.**
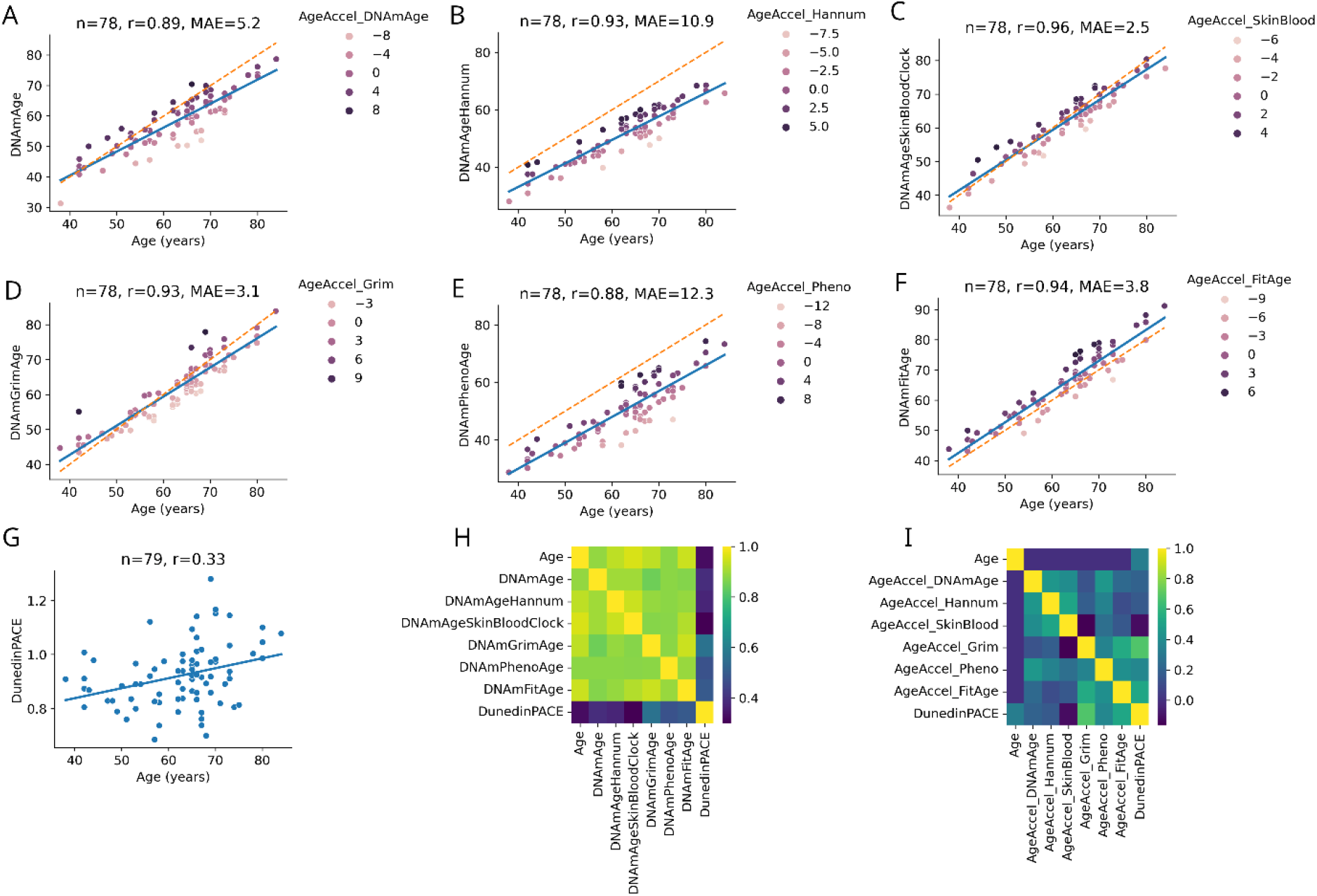
Application of epigenetic clocks in our cohort. (A-G) Predicted age (i.e. epigenetic age) of the six epigenetic clocks as well as pace of aging by DunedinPACE. Number of samples (n), Pearson correlation coefficient (r) and mean absolute error (MAE) are indicated for each clocks. Linear regression line (solid blue lines) of the predicted ages is also shown. Age acceleration (i.e. the deviation from the trend) is illustrated by coloring. The dashed orange line is the diameter (x=y). **(H)** Pearson correlations among age and the predicted values of the epigenetic clocks. **(I)** Pearson correlations among age, age accelerations and pace of aging. The significance of correlations is not indicated.

**Extended Data Fig. 3.**
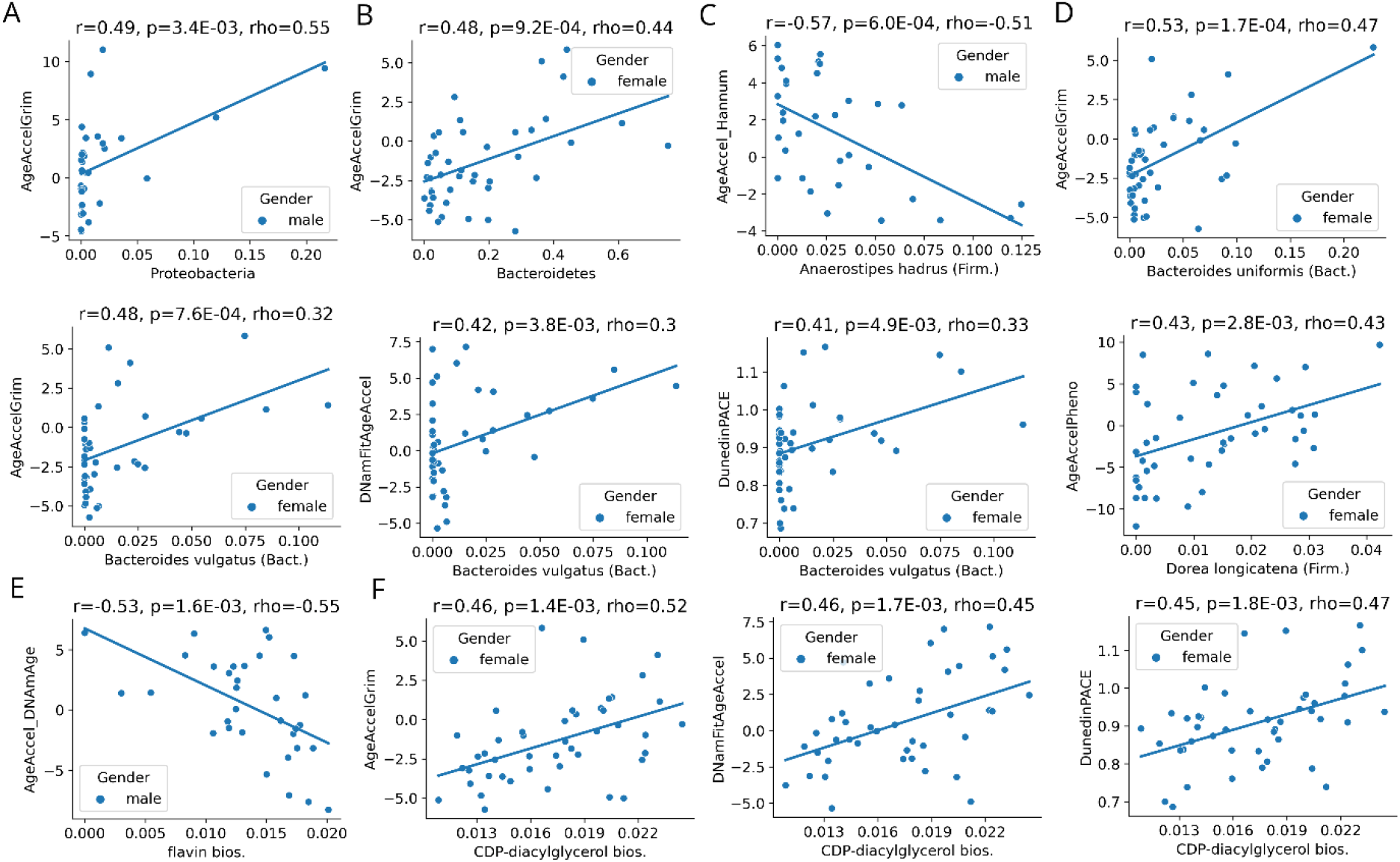
Strongest associations (**, p <= 0.01) between the gut microbiome and epigenetic clocks (associated to Fig. 2). **(A)** Phylum level for males. **(B)** Phylum level for females. **(C)** Species level for males. **(D)** Species level for females. **(E**) Pathway analysis for males. **(F)** Pathway analysis for females. Pearson correlation coefficient (r) the correspondent p-value (p), as well as, Spearman correlation coefficient (rho) are indicated. The regression line (solid blue line) is also shown. Abbreviations are the same as for Fig 2.

**Extended Data Fig. 4.**
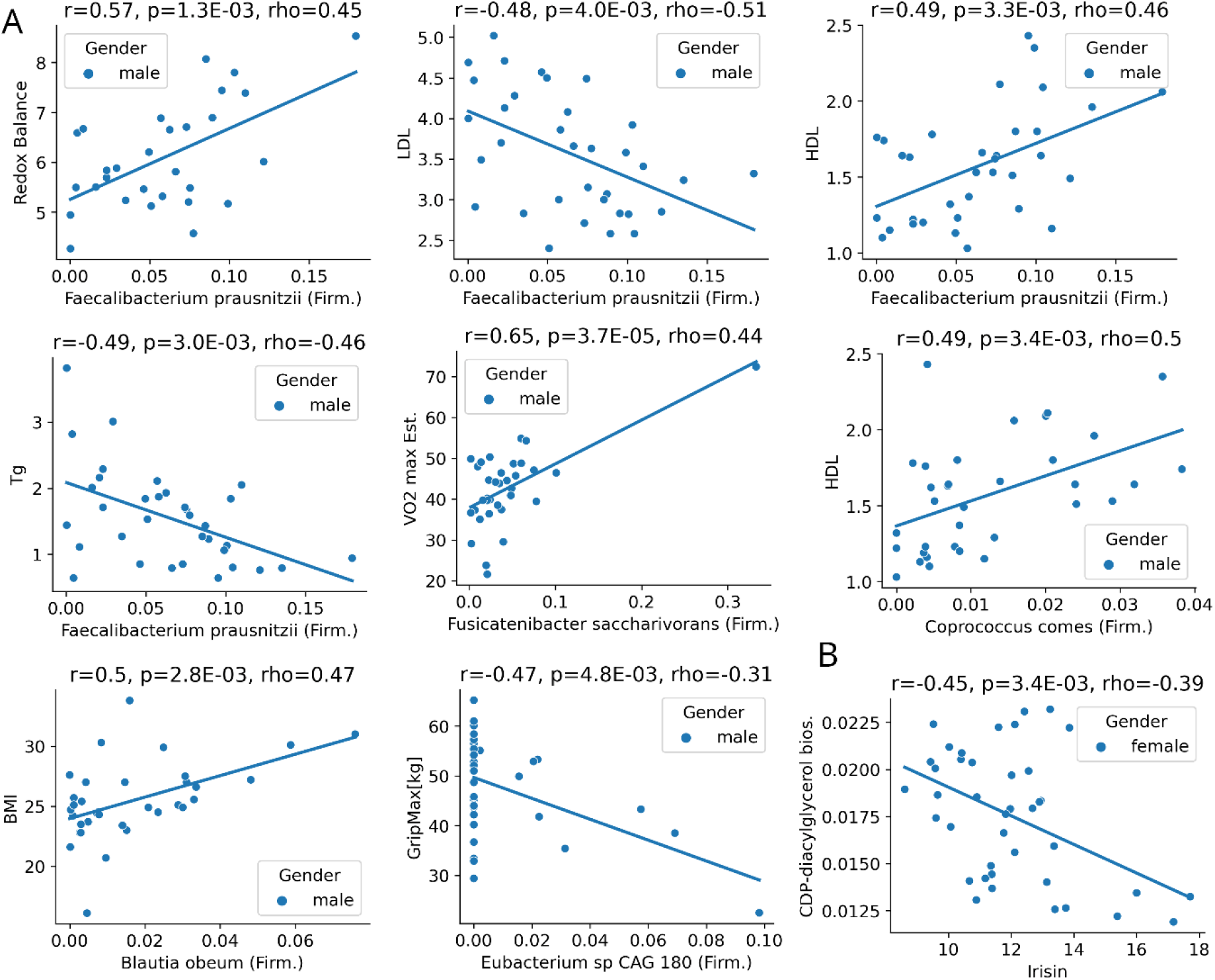
Strongest associations (**, p < 0.01) between the gut microbiome and exercise-related parameters (associated to Fig. 3). **(A)** Species level for males. **(B)** Pathway analysis for females. Pearson correlation coefficient (r), the correspondent p-value (p) as well as Spearman correlation coefficient (rho) are indicated. Regression line (solid blue line) is also shown. Abbreviations is the same as in Fig 3.

